# A Versatile, Spectrophotometer-Based, Quantitative, Visual, Bias-Reducing Method for Human Skin Colour Measurement and Classification

**DOI:** 10.64898/2026.07.27.26358883

**Authors:** Ophelia E. Dadzie, Richard A. Sturm, Sibel Ali, Damilola Fajuyigbe, Antoine Petit, Nina G. Jablonski, George Yu

## Abstract

We evaluated an inexpensive portable hand-held spectrophotometer for skin colour measurement and classification. Under standardised conditions, skin reflectance and colorimetric data were collected from 40 participants of diverse ancestral backgrounds at three anatomical sites: forehead (FH), right posterior forearm (FA), and right upper inner arm (RUA). Demographic and ancestral data, Fitzpatrick Skin Phototype classification, standardised iPhone 13 images, and visual and device-based skin colour matches were also obtained. Participants spanned the five-point EHSCS scale; visually matched Pantone SkinTone colours numbered 25 for FH, 33 for FA, and 28 for RUA. ITA values derived from colorimetric data were used to generate site-specific classifications using the five-point EHSCS, seven-point ITA scale incorporating Del Bino categories, and ten-point MST categories defined by Ulrich or Lipnick. This versatile spectrophotometer-based approach supports affordable, reproducible, race-, ethnicity-, and ancestry-independent skin colour measurement and classification across multiple scales for dermatologists and other users.

## INTRODUCTION

Dermatology’s clinical knowledge base was historically shaped largely by observations in light-skinned populations, limiting attention to pigmentary diversity.^1^ Although recent work has sought to include the full spectrum of human skin pigmentation,^2–3^ progress remains hindered by the absence of precise, universally accepted classification systems and by the inappropriate use of Fitzpatrick skin phototype (FSP), race, ethnicity, or geography as proxies for skin colour.^4–6^ Clinical research and guideline development nevertheless require practical categories within the continuous spectrum of pigmentation. Current proposed frameworks include the Eumelanin Human Skin Colour Scale (EHSCS), based on skin reflectance data (melanin index);^7^ the Individual Typology Angle (ITA), derived from CIE L*a*b* colorimetric parameters;^8–9^ and the Monk Skin Tone Scale (MST), based on visual perception of ten skin-colour representations.^10^ However, there remains a need for a versatile, affordable, reproducible, race-, ethnicity-, and ancestry-independent skin colour measurement method and classification system that will be universally-agreed upon, and of use to diverse constituents, including dermatologists, skin biologists, anthropologists and other users.

The Spectro 1 Pro (Variable Inc)^11^ is an inexpensive, compact, portable hand-held spectrophotometer that measures Hunter L, a, b and CIE L*, a*, b* colour coordinates by spectral analysis across 400–700 nm. It uses the Pantone Colour Matching System (PCMS) for standardized colour reproduction and is widely used in paint and interior design for reproducible colour selection and matching but has not previously been applied to human skin colour measurement.

This study evaluated the utility of this hand-held spectrophotometer for skin colour measurement and classification and assessed its accuracy and reliability for skin colour matching compared with unaided visual assessment. Although this device was used in this study, our proposed methodology is device-agnostic and other spectrophotometers could be utilised.

## MATERIALS AND METHODS

Triplicate skin reflectance and colorimetric measurements were obtained from three anatomical sites (right upper inner arm (RUA), right posterior forearm (FA), and forehead (FH)) in 40 participants of diverse ancestral backgrounds using the spectrophotometer, calibrated daily. At each site, three standardized iPhone 13 images were captured, and skin colour was matched to the Pantone SkinTone Guide (PSTG)^12^ both instrumentally and visually, with reference PSTG images captured using the same device.

All measurements, imaging, and PSTG matching were performed under standardized conditions by one investigator (OED) in a darkened room using only the Sunmatch Scagrip 4, positioned 1 metre from the participant and set at 5500 Kelvins.

Participant data included age, sex, self-identified ethnic group (2021 Census England and Wales), country of birth for participants, parents and grandparents, self- and investigator-classified FSP, hair colour at age 21 years, eye colour, height, weight, and medical and medication history.

Participants were recruited in London, United Kingdom, via a public study website, including individuals referred by a market research company.

**Figure 1** presents an overview of the experimental schedule, including the inclusion and exclusion criteria. The comprehensive data collection sheet is available in Supplementary Material 1.

**Figure 1.**
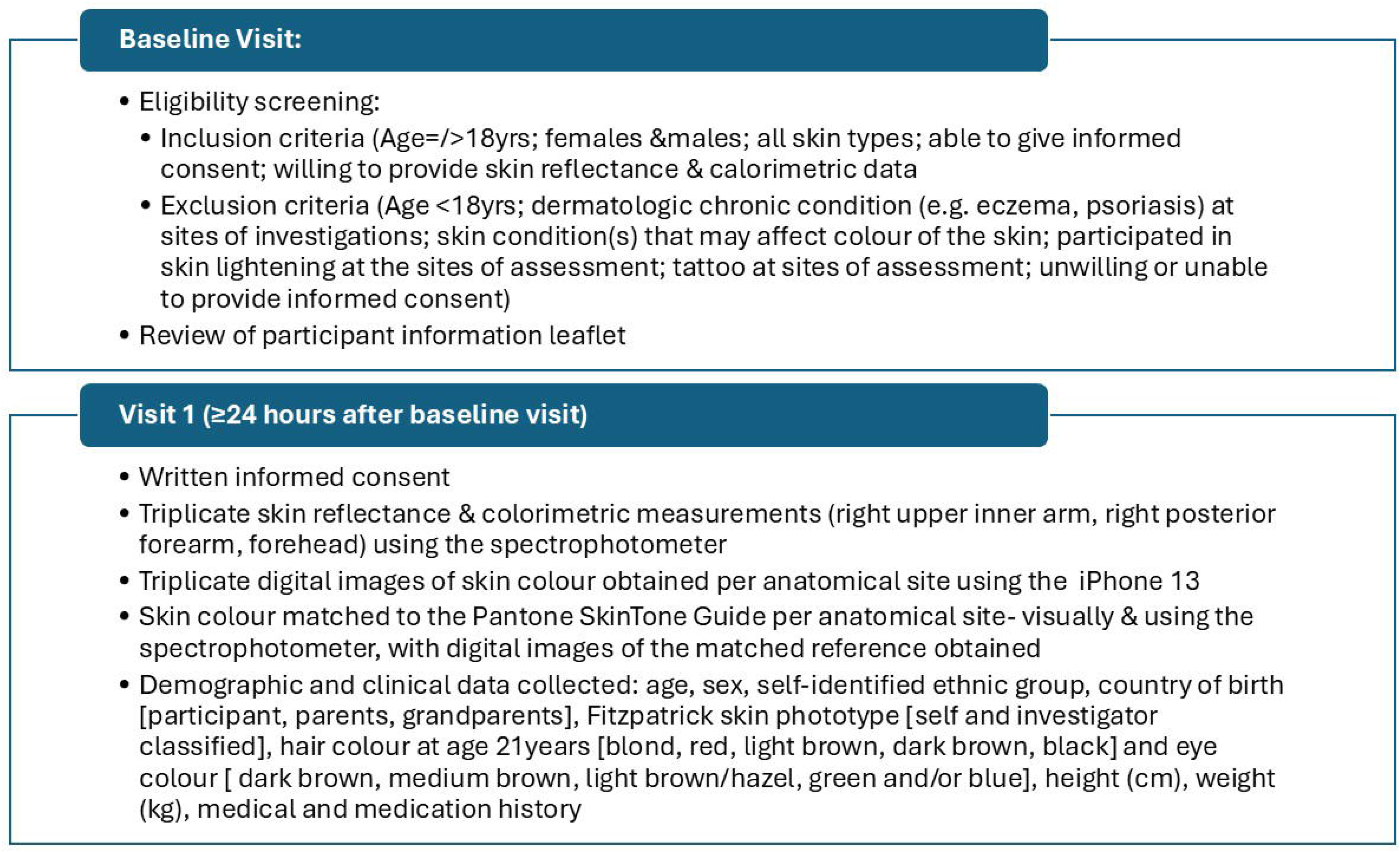
Overview of Experimental Schedule.

## RESULTS

### Pigmentation phenotypes of participants selected to study a range of human skin colours

Forty London-based participants were recruited to span the five-point EHSCS human skin colour scale (**Table 1**). The cohort was 57.5% female, with a mean age of 46.8 ± 14.5 years and all hair colours except red represented. Participant-reported ancestry and ethnic group (2021 Census England and Wales; **Table S1**) were pooled as Black (37.5%), White (32.5%), Asian (20%), and Mixed/Middle Eastern (10%). This distribution was broadly comparable to a recent skin pigment diversity study using NIH race categories.^13^ Investigator-assessed FSP ranged from 1 (5%) to 6 (20%) and broadly aligned with self-classification.

**Table 1.**
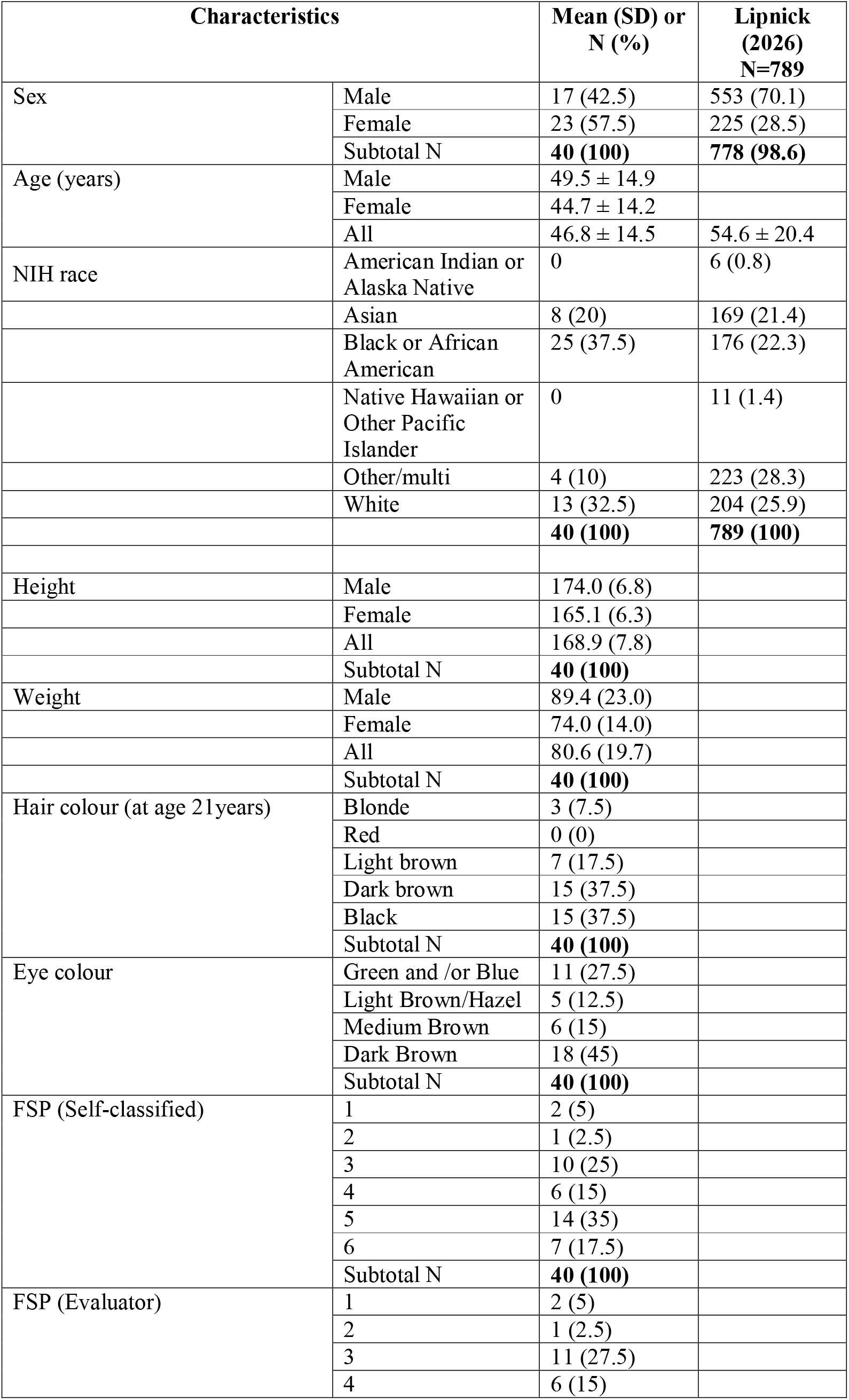
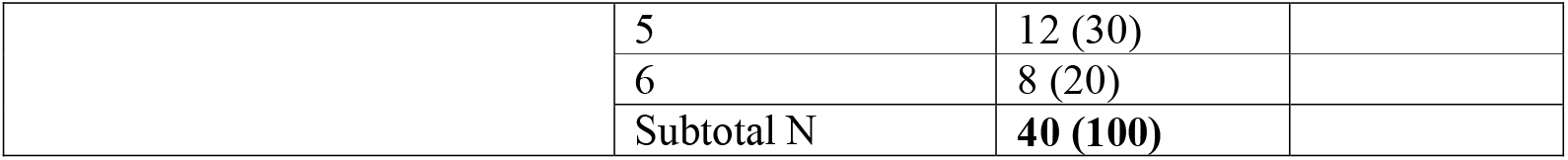
Phenotypic features of 40 patients examined for skin colour.

| Characteristics |  | Mean (SD) or<br>N (%) | Lipnick<br>(2026)<br>N=789 |
| --- | --- | --- | --- |
| Sex | Male | 17 (42.5) | 553 (70.1) |
|  | Female | 23 (57.5) | 225 (28.5) |
|  | Subtotal N | **40 (100)** | **778 (98.6)** |
| Age (years) | Male | 49.5 ± 14.9 |  |
|  | Female | 44.7 ± 14.2 |  |
|  | All | 46.8 ± 14.5 | 54.6 ± 20.4 |
| NIH race | American Indian or<br>Alaska Native | 0 | 6 (0.8) |
|  | Asian | 8 (20) | 169 (21.4) |
|  | Black or African<br>American | 25 (37.5) | 176 (22.3) |
|  | Native Hawaiian or<br>Other Pacific<br>Islander | 0 | 11 (1.4) |
|  | Other/multi | 4 (10) | 223 (28.3) |
|  | White | 13 (32.5) | 204 (25.9) |
|  |  | **40 (100)** | **789 (100)** |
| Height | Male | 174.0 (6.8) |  |
|  | Female | 165.1 (6.3) |  |
|  | All | 168.9 (7.8) |  |
|  | Subtotal N | **40 (100)** |  |
| Weight | Male | 89.4 (23.0) |  |
|  | Female | 74.0 (14.0) |  |
|  | All | 80.6 (19.7) |  |
|  | Subtotal N | **40 (100)** |  |
| Hair colour (at age 21years) | Blonde | 3 (7.5) |  |
|  | Red | 0 (0) |  |
|  | Light brown | 7 (17.5) |  |
|  | Dark brown | 15 (37.5) |  |
|  | Black | 15 (37.5) |  |
|  | Subtotal N | **40 (100)** |  |
| Eye colour | Green and /or Blue | 11 (27.5) |  |
|  | Light Brown/Hazel | 5 (12.5) |  |
|  | Medium Brown | 6 (15) |  |
|  | Dark Brown | 18 (45) |  |
|  | Subtotal N | **40 (100)** |  |
| FSP (Self-classified) | 1 | 2 (5) |  |
|  | 2 | 1 (2.5) |  |
|  | 3 | 10 (25) |  |
|  | 4 | 6 (15) |  |
|  | 5 | 14 (35) |  |
|  | 6 | 7 (17.5) |  |
|  | Subtotal N | **40 (100)** |  |
| FSP (Evaluator) | 1 | 2 (5) |  |
|  | 2 | 1 (2.5) |  |
|  | 3 | 11 (27.5) |  |
|  | 4 | 6 (15) |  |
|  | 5 | 12 (30) |  |
|  | 6 | 8 (20) |  |
|  | Subtotal N | **40 (100)** |  |

### Photographic assessment of skin pigmentation, spectrophotometric measurement and correlation with Pantone Colour Matching System

Triplicate iPhone 13 photographs and spectrophotometric measurements were obtained at FH, FA and RUA under standardised lighting. The spectrophotometer recorded CIE L*, a* and b* values across 400–700 nm.

Because skin translucency may affect instrument-based readings, the investigator (OED) also visually matched skin colour at the same sites using the PSTG, with corresponding images captured for each match.

Visually matched Pantone SkinTone colours totalled 25 for FH, 33 for FA, and 28 for RUA (**Table 2, Table S1 and Figure 2**).

**Figure 2.** Visual comparison of photographic reproduction of skin colour, pantone colour palette of EHSCS types. ***Figure not available in the medRxiv version.*** *Please contact the corresponding author to request access*.

**Table 2.**
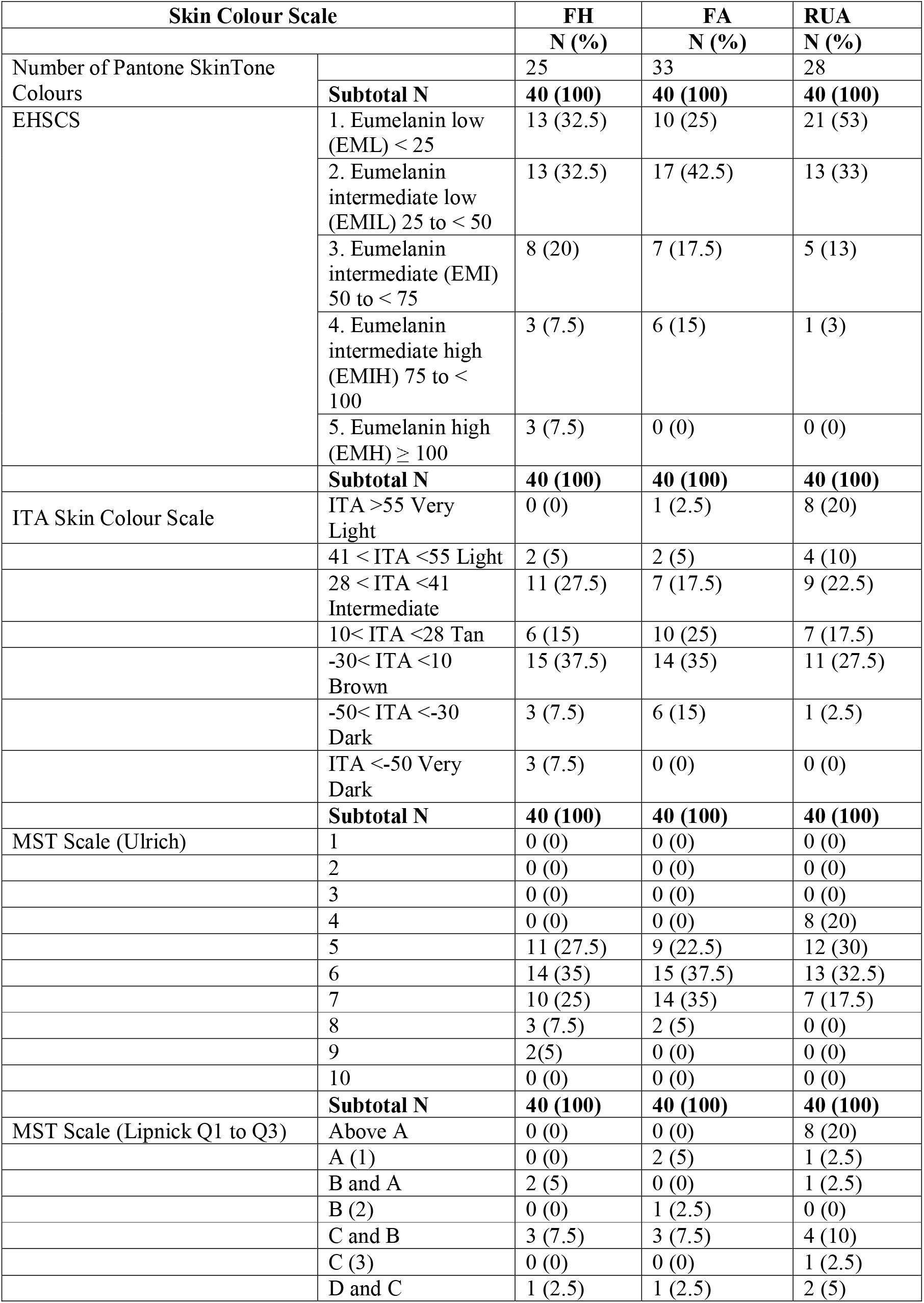
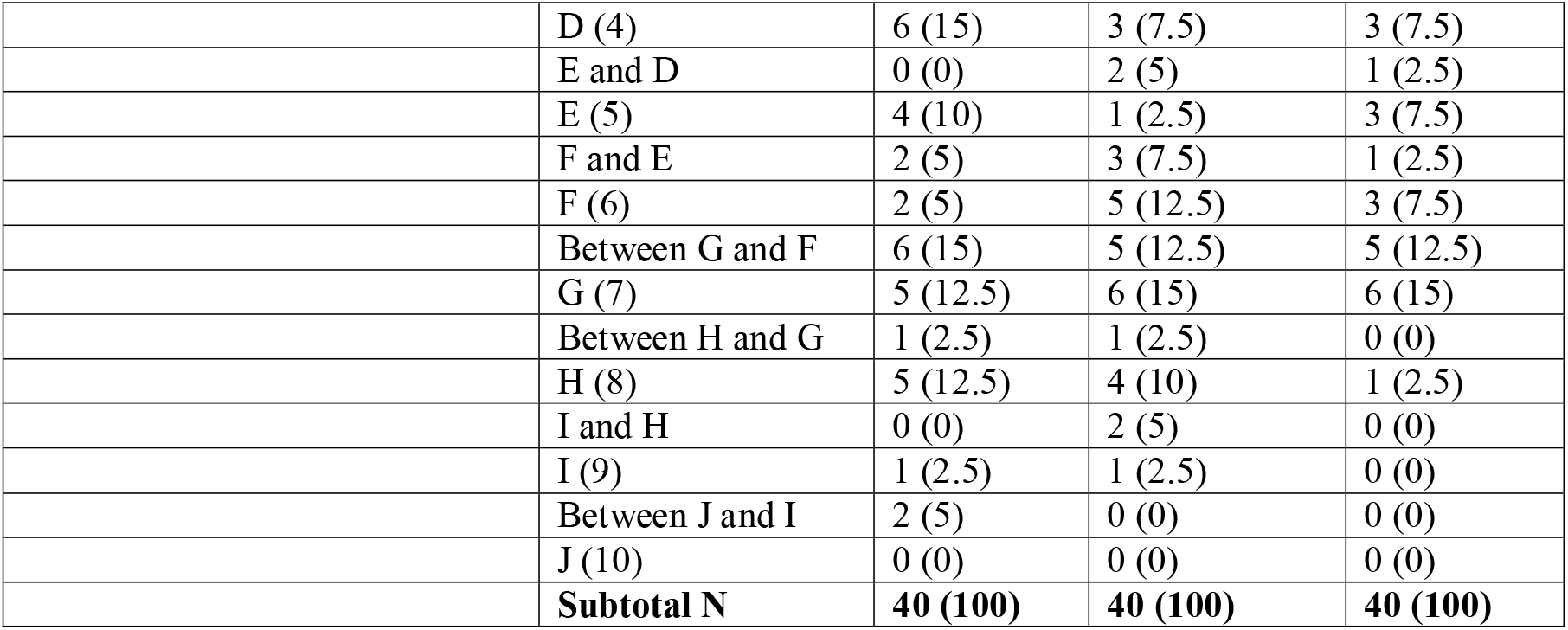
Skin colour scales of the FH, FA and RUA sites for 40 patients examined by spectrophotometric measurement. The number (N) and percent of 40 participants with Pantone SkinTone colours, EHSCS 1 to 5, the seven ITA skin colours groupings, and Monk Skin Tone Scale as 1 to 10 (10 divisions, ^16^) and between A-J (19 designations Q1 to Q3 as per ^13^) are listed at three anatomical locations.

| Skin Colour Scale |  | FH | FA | RUA |
| --- | --- | --- | --- | --- |
|  |  | N (%) | N (%) | N (%) |
| Number of Pantone SkinTone Colours |  | 25 | 33 | 28 |
|  | **Subtotal N** | **40 (100)** | **40 (100)** | **40 (100)** |
| EHSCS | 1. Eumelanin low (EML) < 25 | 13 (32.5) | 10 (25) | 21 (53) |
|  | 2. Eumelanin intermediate low (EMIL) 25 to < 50 | 13 (32.5) | 17 (42.5) | 13 (33) |
|  | 3. Eumelanin intermediate (EMI) 50 to < 75 | 8 (20) | 7 (17.5) | 5 (13) |
|  | 4. Eumelanin intermediate high (EMIH) 75 to < 100 | 3 (7.5) | 6 (15) | 1 (3) |
|  | 5. Eumelanin high (EMH) ≥ 100 | 3 (7.5) | 0 (0) | 0 (0) |
|  | **Subtotal N** | **40 (100)** | **40 (100)** | **40 (100)** |
| ITA Skin Colour Scale | ITA >55 Very Light | 0 (0) | 1 (2.5) | 8 (20) |
|  | 41 < ITA <55 Light | 2 (5) | 2 (5) | 4 (10) |
|  | 28 < ITA <41 Intermediate | 11 (27.5) | 7 (17.5) | 9 (22.5) |
|  | 10 < ITA <28 Tan | 6 (15) | 10 (25) | 7 (17.5) |
|  | -30 < ITA <10 Brown | 15 (37.5) | 14 (35) | 11 (27.5) |
|  | -50 < ITA <-30 Dark | 3 (7.5) | 6 (15) | 1 (2.5) |
|  | ITA <-50 Very Dark | 3 (7.5) | 0 (0) | 0 (0) |
|  | **Subtotal N** | **40 (100)** | **40 (100)** | **40 (100)** |
| MST Scale (Ulrich) | 1 | 0 (0) | 0 (0) | 0 (0) |
|  | 2 | 0 (0) | 0 (0) | 0 (0) |
|  | 3 | 0 (0) | 0 (0) | 0 (0) |
|  | 4 | 0 (0) | 0 (0) | 8 (20) |
|  | 5 | 11 (27.5) | 9 (22.5) | 12 (30) |
|  | 6 | 14 (35) | 15 (37.5) | 13 (32.5) |
|  | 7 | 10 (25) | 14 (35) | 7 (17.5) |
|  | 8 | 3 (7.5) | 2 (5) | 0 (0) |
|  | 9 | 2 (5) | 0 (0) | 0 (0) |
|  | 10 | 0 (0) | 0 (0) | 0 (0) |
|  | **Subtotal N** | **40 (100)** | **40 (100)** | **40 (100)** |
| MST Scale (Lipnick Q1 to Q3) | Above A | 0 (0) | 0 (0) | 8 (20) |
|  | A (1) | 0 (0) | 2 (5) | 1 (2.5) |
|  | B and A | 2 (5) | 0 (0) | 1 (2.5) |
|  | B (2) | 0 (0) | 1 (2.5) | 0 (0) |
|  | C and B | 3 (7.5) | 3 (7.5) | 4 (10) |
|  | C (3) | 0 (0) | 0 (0) | 1 (2.5) |
|  | D and C | 1 (2.5) | 1 (2.5) | 2 (5) |
|  | D (4) | 6 (15) | 3 (7.5) | 3 (7.5) |
|  | E and D | 0 (0) | 2 (5) | 1 (2.5) |
|  | E (5) | 4 (10) | 1 (2.5) | 3 (7.5) |
|  | F and E | 2 (5) | 3 (7.5) | 1 (2.5) |
|  | F (6) | 2 (5) | 5 (12.5) | 3 (7.5) |
|  | Between G and F | 6 (15) | 5 (12.5) | 5 (12.5) |
|  | G (7) | 5 (12.5) | 6 (15) | 6 (15) |
|  | Between H and G | 1 (2.5) | 1 (2.5) | 0 (0) |
|  | H (8) | 5 (12.5) | 4 (10) | 1 (2.5) |
|  | I and H | 0 (0) | 2 (5) | 0 (0) |
|  | I (9) | 1 (2.5) | 1 (2.5) | 0 (0) |
|  | Between J and I | 2 (5) | 0 (0) | 0 (0) |
|  | J (10) | 0 (0) | 0 (0) | 0 (0) |
|  | **Subtotal N** | **40 (100)** | **40 (100)** | **40 (100)** |

### Analysis of EHSCS skin type categories and comparison with ITA and MST scales

Spectrophotometer-derived L* and b* values were used to calculate ITA at all three sites (**Tables S2 to S4**). Although EHSCS was originally based on melanin content,^7^ its approximate linearity with ITA and melanin index^14^ allows expression as ITA values: >28 (EML 1), -10 to <28 (EMIL 2), -30 to <-10 (EMI 3), -50 to <-30 (EMIH 4), and <-50 (EMH 5) (**Figure S1**).

EHSCS-ITA categories varied by site for most participants, with FH generally darker than FA and RUA lightest. FH classifications were EML 1 32.5%, EMIL 2 32.5%, EMI 3 20%, EMIH 4 7.5%, and EMH 5 7.5%; FA classifications were 25%, 42.5%, 17.5%, 15%, and 0%, respectively; and RUA classifications were 53%, 33%, 13%, 3%, and 0%, respectively (**Table 2**). Three participants classified as EMH 5 on FH were EMIH 4 on FA, and two were EMI 3 on RUA (**Tables S2 to S4**). Only 14 participants (35%) had the same EHSCS category across all sites. Of these, 9 were EML 1 (22.5%) and 5 were EMIL 2 (12.5%).

ITA scores were also classified using the seven-point Del Bino scale (very light > 55° > light > 41° > intermediate > 28° > tan > 10° > brown > −30° > dark;^9^ with the addition of very dark ITA < −50°^15^) and the 10-point MST categories were assigned using both Ulrich^16^ and Lipnick^13^ ITA conversion methods (**Tables S2 to S4**). Results by FH, FA and RUA are shown in **Table 2**.

All three methods again showed progressive lightening from FH to FA to RUA. Consistent classification across all three sites occurred in 8 participants (20%) using ITA, 6 (15%) using MST (Ulrich), and 1 (2.5%) using MST (Lipnick).

### Comparison of skin type categories by EHSCS, ITA, MST, FSP and PSTG

Skin colour categories across the five classification systems are summarised in **Table S5** for FH, FA and RUA, sorted by FH EHSCS category. By definition, EML 1 corresponds to Very Light, Light and Intermediate ITA categories, although Very Light occurred only at RUA in this cohort; EMIL 2 corresponds to Tan and Brown, EMI 3 to Brown, EMIH 4 to Dark, and EMH 5 to Very Dark.

Using Ulrich^16^ MST categories, EML 1 corresponded to MST 5–6, EMIL 2 to 6–7, EMI 3 to 7, EMIH 4 to 7–8, and EMH 5 to 9. Using Lipnick^13^ categories, EML 1 corresponded to MST A–E, EMIL 2 to E–G, EMI 3 to G–H, EMIH 4 to H–I, and EMH 5 to I–J.

Self- and evaluator-classified FSP showed broad overlap with EHSCS: EML 1 ranged from FSP 1–5, EMIL 2 from FSP 3–5, and EMI 3, EMIH 4 and EMH 5 from FSP 5–6.

PSTG provided the most granular classification (**Table 2**, **Table S5**), capturing skin-colour subtleties not reflected in the broader EHSCS, ITA, MST and FSP systems.

## DISCUSSION

Race and skin colour are often conflated in biology and medicine, reinforcing implicit bias and health inequities.^1^ Yet race is historically, temporally, and geographically contingent, whereas skin colour is measurable.^17^ Objective, standardised methods that separate skin colour from ancestry, race, and ethnicity may help reduce structural racism and promote health equity in science and medicine.

This study shows that an inexpensive portable hand-held spectrophotometer provides an objective, standardised method for classifying skin colour by EHSCS, ITA and MST, and for matching skin colour to the PSTG across constitutive and facultative pigmentation. This supports its potential as a reproducible tool for dermatological and clinical research. To our knowledge, this is the first evaluation of the Spectro 1 Pro for skin colour classification, although the methodology we have presented here is device-agnostic and is likely to be applicable to other commercially available hand-held spectrophotometers and with technological improvements that are in development.

Although widely used, FSP was developed to assess UVR responses in lightly pigmented skin, not to classify the full spectrum of human skin colour.^18^ It therefore performs poorly for moderate to darker pigmentation.^4,19–20^ In this study, discrepancies between self- and investigator-assessed FSP, particularly in the FSP 2/3 and 5/6 groups, underscored its subjectivity. Participants classified as FSP 5 or 6 also spanned multiple EHSCS categories, confirming that FSP compresses substantial pigmentary variation into too few categories.^21^

Visual assessment also showed limitations, with both over- and under-estimation of pigmentation even by a trained clinician, highlighting the value of objective measurement in reducing visual bias.

EHSCS was the most simplistic scale in this study, providing clear, unambiguous classifications across all participants and anatomical sites while remaining independent of self-reported race, ethnicity, and ancestry.

Although ITA uses continuous colorimetric data and predefined thresholds, its distribution in this cohort appeared skewed toward lighter pigmentation, suggesting stronger discrimination within lighter skin tones. EHSCS may therefore offer more balanced classification across the full pigmentation range.

MST classifications varied by conversion method (Ulrich^16^ vs. Lipnick^13^), indicating limitations in the categorisation framework rather than biological variation. Published evidence on the methodology used to define Monk categories also remains limited.

Skin colour varied by anatomical site, with FH darker than FA and FA darker than RUA, consistent with UVR-related facultative pigmentation in exposed sites and constitutive pigmentation in protected sites. Although pigmentary demarcation lines^22^ may contribute, affected participants would have been excluded. These site-specific differences highlight the spectrophotometer’s sensitivity and support site-specific skin colour measurement.

The main limitation was the small sample size (n=40), although participants represented diverse ancestry, pigmentation, hair and eye colour. Use of a single investigator and clinical setting was also limiting. Larger, multicentre studies with multiple investigators are needed to confirm these findings, assess seasonal pigmentation changes, and evaluate performance under varied conditions.

Objective quantification of cutaneous melanin pigmentation has broad applications in dermatology, skin biology, and related industries. It could support skin-inclusive semiology, improve severity scores reliant on erythema assessment, and refine evaluation of dyschromia, including in Cutaneous Lupus Erythematosus Disease Area and Severity Index (CLASI) where darker skin may lead to underestimation of inflammation and overestimation of pigmentary sequelae.^23–24^ It could also inform studies of post-inflammatory dyschromia, photo-induced malignancy risk, prevention and screening strategies, cosmetic applications, and personalised laser or energy-based device settings.

A further distinguishing feature of our work is the use of several skin colour classification systems within a single methodology, rather than the single-system approach typical of prior studies. This reflects our view that no one system is likely to suit every clinical or research need: the optimal choice depends on the question being asked, and in some cases a combination of systems may offer the greatest insight. Rather than advocating for a single “best” classification system, this approach offers the flexibility to generate and apply multiple complementary systems as appropriate to the context.

In summary, we present a versatile, affordable, reproducible, spectrophotometer-based method for skin colour measurement and classification that is race-, ethnicity-, and ancestry-independent, allowing objective and standardised skin colour classification across multiple scales. Employing such a technique would enable reliable and reproducible measurement, description and communication about skin colour by dermatologists and other users, with potential to promote equity in medicine and science.

## Declaration of Generative Artificial Intelligence (AI) or Large Language Models (LLMs)

**The author(s) did not use AI/LLM in any part of the research process and/or manuscript preparation**

## Human Ethics

Ethics approval and consent to participate: Ethics approval was granted by HRA and Health and Care Research Wales (HCRW) and informed consent regarding future use of samples for research was signed for by all participants contributing biospecimens. Consent for publication: All participants contributing biospecimens to this study gave informed consent for publication of data generated using their samples.

## Study Site/Research Governance Statement

The study was conducted at an independent private research facility (Parexel International, Northwick Park Hospital, Watford Rd, Harrow HA1 3UJ).

## Data Availability

The data underlying this article are available in the article and in its online supplementary material.

## Acknowledgements

We thank Mr Christian Dietz Managing Director RIG/GmbH/Director of Technology Rhopoint Instruments Ltd for technical advice on colour matching principles.

## AUTHOR CONTRIBUTIONS

Conceptualization (OED, RAS, AP, NGJ and GY); Data Curation (OED, RAS and GY); Formal Analysis (RAS, OED, SA and GY); Funding Acquisition (OED, NGJ); Investigation (OED); Methodology (OED, RAS, DF, AP, NGJ, GY); Project Administration (OED); Resources (OED, RAS, DF and GY); Software (OED, RAS, GY); Supervision (OED); Validation (OED, and RAS), Visualization: (OED, RAS, SA and GY); Writing - Original Draft Preparation (OED, RAS and NGJ); Writing - Review and Editing (OED, RAS, SA, DF, AP, NGJ and GY)

## List of Abbreviations

EHSCS: Eumelanin Human Skin Colour Scale
FA: Right Posterior Forearm
FH: Forehead
FSP: Fitzpatrick Skin Phototype
MST: Monk Skin Tone Scale
PSTG: Pantone SkinTone Guide
RUA: Right Upper Inner Arm

---

**Figure S1**. Individual typology angle (ITA) and melanin index (MI) in an African population, correlated with the Eumelanin Human Skin Colour Scale (EHSCS). The figure is based on the EHSCS scale as represented as a plot of ITA and MI in an African population (Dadzie et al., 2022)^7^. The EHSCS quantiles 1 to 5 at the top and right-hand side of the plot, and with vertical and horizontal lines in colour (lightest to darkest) indicating the divisions, the corresponding categories are shown at the bottom of the plot as MI scores and on the left-hand side as ITA values.

**Table S1.** Phenotypic characteristics of 40 patients and Pantone Colour Matching System.

***Table S1 not available in the medRxiv version.*** *Please contact the corresponding author to request access*.

**Table S2.** Skin colour type on FH for 40 patients on EHSCS, ITA and MST scales

Thesubject and measured position is shown in Column A and the calculated ITA score using the spectrophotometer L* and b* mean values from three readings is shown in Column B. The ITA to EHSCS conversion and the corresponding EHSCS scale is shown in Column C and D, the ITA skin colour scale value conversion and ITA scale are shown in Columns E and F. The ITA to MST (Ulrich)^16^ conversion and MST scale are shown in Columns G and H, with ITA to MST (Lipnick Q1 to Q3)^13^ conversion and MST scale are shown in Columns I and J.

**Table S3.** Skin colour type on FA for 40 patients on EHSCS, ITA and MST scales Columns are as per Table S2.

**Table S4.** Skin colour type on RUA for 40 patients on EHSCS, ITA and MST scales Columns are as per Table S2.

**Table S5**. Skin colour type on FH, FA and RUA for 40 patients based on EHSCS, ITA, MST, FSP and Pantone colour scales.

The subject ID is shown in Column A with the calculated ITA score using the spectrophotometer for FH, FA, RUA sites in Columns B to D. The scores for EHSCS, ITA, MST (Ulrich^16^ and Lipnick^13^), FSP (Self and Evaluator classifications), and Pantone SkinTone are shown for FH (Columns F to L), FA (Columns N to T), RUA (Columns V to AB), are shown across the Table respectively. The skin type classifications are sorted on EHSCS for the FH (Column F).

## REFERENCES

1. Horsley V, Dadzie OE, Hall R, Jablonski NG. Disentangling race from skin colour in modern biology and medicine. J Invest Dermatol 2025; 145: 240–248.

2. Takeshita T, McMichael AJ, Miller-Monthrope Y et al. Rethinking the use of population descriptors in dermatology trials and beyond: disentangling race and ethnicity from skin color. Arch Dermatol Res 2025; 317: 728.

3. Burgess CM, Byrd AS, Cobb CBC et al. A call to action. The need to update population descriptors in dermatology research studies with the use of an inclusive skin classification system. J Am Acad Dermatol 2025; 93: 553–556.

4. Eilers S, Bach DQ, Gaber R et al. Accuracy of self-report in assessing Fitzpatrick skin phototypes I through VI. JAMA Dermatol 2013; 149: 1289–1294.

5. Harvey VM, Alexis A, Okeke CAV et al. Integrating skin color assessments into clinical practice and research: a review of current approaches. J Am Acad Dermatol 2024; 91: 1189–1198.

6. Sarkar R, Mehta H. A pigment-informed dermatology framework — reimagining “Skin of Color”. JAMA Dermatol 2026; 162: 441–442.

7. Dadzie OE, Sturm RA, Fajuyigbe D et al. The Eumelanin Human Skin Colour Scale: a proof-of-concept study. Br J Dermatol 2022; 187: 99–104.

8. Chardon A, Cretois I, Hourseau C. Skin colour typology and suntanning pathways. Int J Cosmet Sci 1991; 13: 191–208.

9. Del Bino S, Bernerd F. Variations in skin colour and the biological consequences of ultraviolet radiation exposure. Br J Dermatol 2013; 169 (Suppl. 3): 33–40.

10. Monk E. Monk Skin Tone Scale [WWW Document]. 2019. URL https://skintone.google [accessed on 10 June 2026].

11. Variable Inc. Spectro 1 Pro: Affordable Precision Spectrophotometer [WWW Document].URLhttps://variableinc.com/product/spectro-1-pro-affordable-precision-spectrophotometer/ [accessed on 10 June 2026].

12. Pantone. PANTONE SkinTone Guide [WWW Document]. URL https://www.pantone.com/uk/en/skintone [accessed on 13 June 2026].

13. Lipnick MS, Chen D, Law T et al. Comparison of methods for characterizing skin pigment diversity in research cohorts. Br J Dermatol 2026; 194: 135–145.

14. Wilkes M, Wright CY, du Plessis JL et al. Fitzpatrick Skin Type, Individual Typology Angle, and Melanin Index in an African population: steps toward universally applicable skin photosensitivity assessments. JAMA Dermatol 2015; 151: 902–903.

15. Rutjes C, Primiero CA, Mothershaw A et al. Automating skin colour assessment using 3D total-body photography: a proof-of-concept study. J Eur Acad Dermatol Venereol 2026; 40: 528–530.

16. Ulrich P, Zink A, Biedermann T, et al. Beyond Fitzpatrick: automated artificial intelligence-based skin tone analysis in dermatological patients. NPJ Digit Med 2025; 8: 378.

17. Jablonski NG. Skin color and race. Am J Phys Anthropol 2021; 175: 437–447.

18. Fitzpatrick TB. Soleil et peau [Sun and skin]. J Méd Esthétique 1975; 2: 33–34.

19. Fitzpatrick TB. The validity and practicality of sun-reactive skin types I through VI. Arch Dermatol 1988; 124: 869–871.

20. Okoji UK, Taylor SC, Lipoff JB. Equity in skin typing: why it is time to replace the Fitzpatrick scale. Br J Dermatol 2021; 185: 198–199.

21. He SY, McCulloch CE, Boscardin WJ et al. Self-reported pigmentary phenotypes and race are significant but incomplete predictors of Fitzpatrick skin phototype in an ethnically diverse population. J Am Acad Dermatol 2014; 71: 731–737.

22. James WD, Carter JM, Rodman OG. Pigmentary demarcation lines: a population survey. J Am Acad Dermatol 1987; 16: 584–590.

23. McMichael A, Frey C. Challenging the tools used to measure cutaneous lupus severity in patients of all skin types. JAMA Dermatol 2025; 161: 9–10.

24. Xie L, Faden DF, Stone CJ et al. Subtype and racial erythema variation for cutaneous lupus trials. JAMA Dermatol 2025; 161: 67–74.

