## Supplementary figures and images for "A Versatile, Spectrophotometer-Based, Quantitative, Visual, Bias-Reducing Method for Human Skin Colour Measurement and Classification"

### Supplementary Figure 1

#### Slide 1
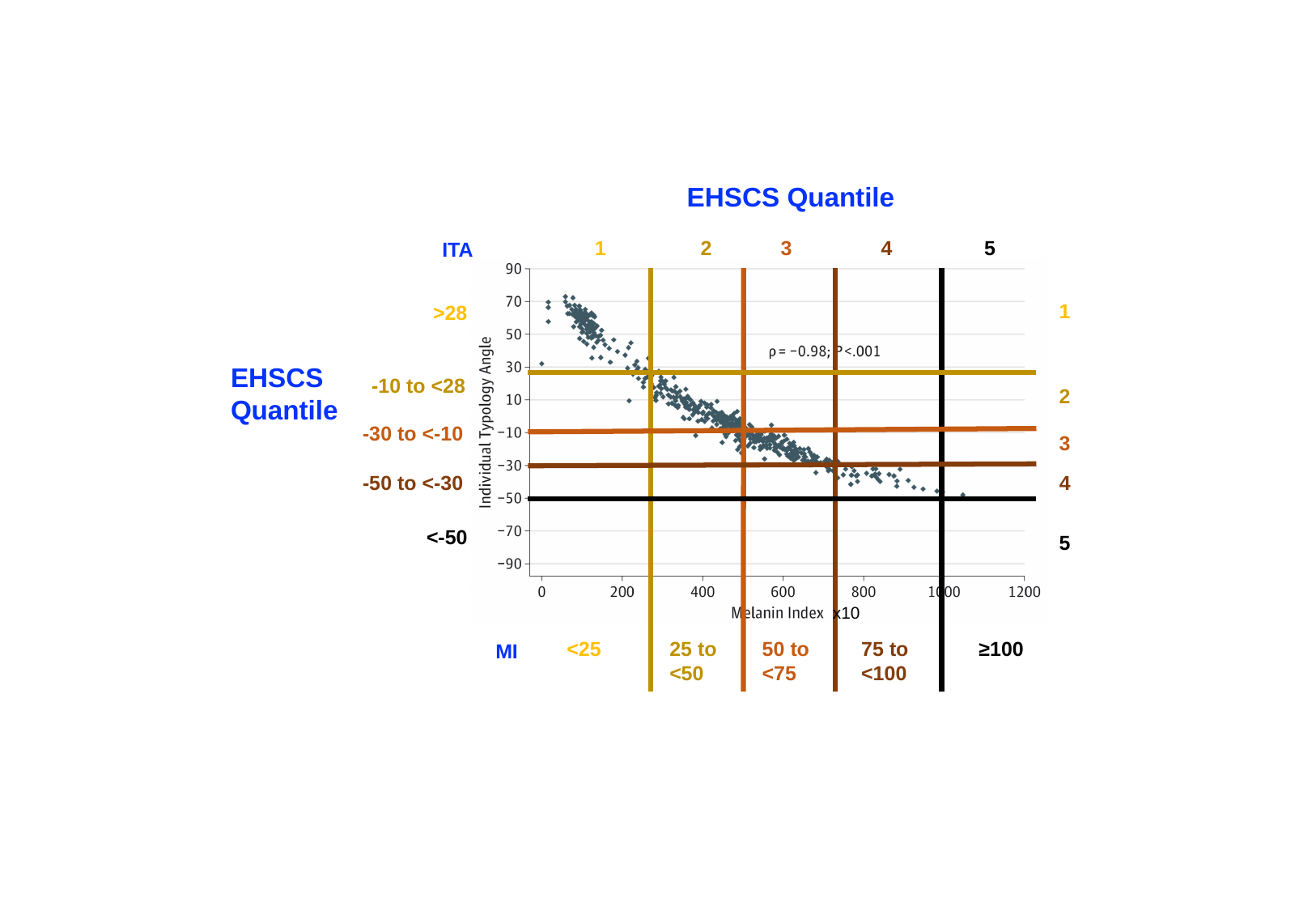

EHSCS Quantile
1
2
3
4
5
ITA
1
>28
EHSCS Quantile
-10 to <28
2
-30 to <-10
3
-50 to <-30
4
<-50
5
x10
<25
25 to <50
50 to <75
75 to <100
≥100
MI
