## Supplementary Material for "A Versatile, Spectrophotometer-Based, Quantitative, Visual, Bias-Reducing Method for Human Skin Colour Measurement and Classification"

**The Eumelanin Human Skin Colour Scale (CRF)**
**Version 1**
**Date: 2nd April 2024**

Subject No:

Date of evaluation:

Location (Location Co-ordinates):

Age:

Sex:

Country of Birth:

Country of Birth of biological Father (if known):

Country of birth of biological mother (if known):

Country of birth of paternal grand-father (if known):

Country of birth of paternal grand-mother (if known):

Country of birth of maternal grand-father (if known):

Country of birth of maternal grand-mother (if known):

**The Eumelanin Human Skin Colour Scale (CRF)**
**Version 1**
**Date: 2nd April 2024**

Subject No:

Date of evaluation:

Location (Location Co-ordinates):

Self-identified ethnic group (using categories for 2021 Census England and Wales):

| | |
|---|---|
| **Asian or Asian British** | Indian |
| | Pakistani |
| | Bangladeshi |
| | Chinese |
| | Any other Asian background |
| **Black, Black British, Caribbean or African** | Caribbean |
| | African |
| | Any other Black, Clack British or Caribbean background |
| **Mixed or multiple ethnic groups** | White and Black Caribbean |
| | White and Black African |
| | White and Asian |
| | Any other Mixed or multiple ethnic background |
| **White** | English, Welsh, Scottish, Northern Irish or British |
| | Irish |
| | Gypsy or Irish Traveller |
| | Roma |
| | Any other White Background |
| **Other Ethnic Group** | Arab |
| | Any other Ethnic Group |

Self-classified Fitzpatrick Skin Phototype:

| Skin Type | Tanning ability |
|---|---|
| I | Always burns, does not tan |
| II | Burns easily, tans poorly |
| III | Tans after initial burn |
| IV | Burns minimally, tans easily |
| V | Rarely burns, tans darkly easily |
| VI | Never burns, always tans darkly |

**The Eumelanin Human Skin Colour Scale (CRF)**
**Version 1**
**Date: 2nd April 2024**

Subject No:

Date of evaluation:

Location (Location Co-ordinates):

Past Medical History (in particular any disorders of pigmentation e.g. vitiligo):

Drug History:

Eye Colour:

Dark brown

Medium brown

Light brown /hazel

Green and/or blue

Hair Colour (at age 21)

Blonde

Red

Light Brown

Dark Brown

Black

**The Eumelanin Human Skin Colour Scale (CRF)**
**Version 1**
**Date: 2nd April 2024**

Subject No:

Date of evaluation:

Location (Location Co-ordinates):

Fitzpatrick Skin Phototype Classified by Evaluator:

| Skin Type | Tanning ability |
|---|---|
| I | Always burns, does not tan |
| II | Burns easily, tans poorly |
| III | Tans after initial burn |
| IV | Burns minimally, tans easily |
| V | Rarely burns, tans darkly easily |
| VI | Never burns, always tans darkly |

Blood pressure:

Height:

Weight:

Pantone SkinTone Guide Colour:

Right upper inner arm

Right forearm

Forehead
